# Comparison of dried blood spots and venous blood for the detection of SARS-CoV-2 antibodies in a population of nursing home residents

**DOI:** 10.1101/2021.05.17.21256410

**Authors:** Eline Meyers, Stefan Heytens, Asangwing Formukong, Hanne Vercruysse, An De Sutter, Tom Geens, Kenneth Hofkens, Heidi Janssens, Eveline Nys, Elizaveta Padalko, Ellen Deschepper, Piet Cools

**Affiliations:** Department of Diagnostic Sciences, Faculty of Medicine and Health Sciences, Ghent University, Ghent, Belgium; Department of Public Health and Primary Care, Faculty of Medicine and Health Sciences, Ghent University, Ghent, Belgium; Research and analytics, Liantis Occupational Health Services, Bruges, Belgium; Laboratory of Medical Microbiology, Ghent University Hospital, Ghent, Belgium; Biostatistics Unit, Faculty of Medicine and Health Sciences, Ghent University, Ghent, Belgium

**Author notes:** Both authors contributed equally to this work (Eline Meyers, Stefan Heytens). Author order was determined in order of increasing seniority. Corresponding author: Prof. Dr. Piet Cools.

**Keywords:** Elderly, nursing homes, SARS-CoV-2, COVID-19, antibodies, immunoglobins, IgG, serology, dried blood spots (DBS), enzyme-linked immunosorbent assay (ELISA), serosurveillance, vaccination

## Abstract

**Introduction:** In the current SARS-CoV-2 pandemic, testing for SARS-CoV-2 specific antibodies is paramount to monitor immune responses in post-authorization vaccination and sero-epidemiology studies. However, large scale and iterative serological testing by venipuncture in older persons can be challenging. Capillary blood sampled using a finger prick and collected on protein saver cards, i.e., dried blood spots (DBS), has already proven to be a promising alternative. However, elderly persons have a reduced cutaneous microvasculature, which may affect DBS-based antibody testing. Therefore, we aimed to evaluate the performance of DBS for the detection of SARS-CoV-2 antibodies in nursing homes residents.

**Materials and methods:** We collected venous blood and paired Whatman and EUROIMMUN DBS from nursing home residents, and from staff as a reference population. Venous blood samples were analyzed for the presence of SARS-CoV-2 IgG antibodies using the Abbot chemiluminescent microparticle immunoassay (CMIA). DBS were analyzed by the EUROIMMUN enzyme-linked immuno sorbent assay (ELISA) for SARS-CoV-2 IgG antibodies. We performed a statistical assessment to optimize the ELISA cut-off value for the DBS using the Youden’s J index.

**Results:** A total of 273 paired DBS-serum samples were analyzed, of which 129 were positive as assessed by the reference test. The sensitivities and specificities of DBS ranged from 95.0% to 97.1% and from 97.1% to 98.8%, respectively, depending on population (residents or staff) or DBS card type.

**Conclusion:** DBS sampling is a valid alternative to venipuncture for the detection of SARS-CoV-2 antibodies in the elderly.

## Introduction

In the current severe acute respiratory syndrome-coronavirus-2 (SARS-CoV-2) pandemic, large-scale serological antibody studies are paramount to assess the true SARS-CoV-2 infection rate.

Indeed, statistics on PCR-confirmed SARS-CoV-2 cases are far from ideal to estimate the true proportion of the population that experienced a SARS-CoV-2 infection (1), as mildly affected or asymptomatic individuals are often not tested and PCR-based testing only yields an epidemiological snapshot. Furthermore, the recent implementation of newly developed SARS-CoV-2 vaccines creates an urgent need for population-based serological studies to monitor antibody responses post-authorization. Protocols for SARS-CoV-2 antibody assays using serum/plasma obtained by venipuncture are well established in a clinical setting. However, venipuncture is invasive and can cause serious discomfort. Especially in the elderly, venipuncture poses large challenges due to dehydration, loss of vein patency and low blood pressure. Elderly may also suffer from arthritis, injury or stroke, impeding to hyperextend the arms to survey for available veins. The use of venipuncture is further limited for wider application in non-clinical settings due to the costs (for e.g., phlebotomists), logistical constraints associated with collecting, processing, and transporting venous blood.

Collecting capillary blood on protein saver cards after finger pricking using a lancet, the so-called dried blood spot (DBS), is a most valuable alternative as it is minimally invasive, can be done at low cost, has the potential for self-sampling and is easy to ship and store. DBS are increasingly applied as a minimally invasive alternative for infectious serology, especially in community- and population-based epidemiology, including SARS-CoV-2 (2). However, the use of DBS has not yet been validated for detection of SARS-CoV-2 antibodies in the elderly. Aging is known to modify the cutaneous microvasculature and the structures of blood vessels, even down to the level of the capillary basement membrane (3). Here, we aimed to validate the use of DBS sampling for the detection of SARS-CoV-2 IgG antibodies in a population of residents and staff from nursing homes.

## Methodology

### Ethical considerations

The current study was approved by the Ethical Committee of the Ghent University Hospital (reference number BC-07665) and conducted according to the principles outlined in the Declaration of Helsinki. Each participant signed an informed consent form after being informed about the goal of the study and the sampling procedures. A confidential counselor, such as a nurse, signed for participants who were incapable to sign the consent form, such as residents with dementia when consent was given by their legal representative.

### Study population

In August 2020, we contacted the management of four NHs within our network (Amphora, Wingene; Sint-Rafaël, Liedekerke; Sint-Jozef, Assenede and Sint-Jozef, Wetteren; all in Flanders, Belgium) and explained the goal of the study. We recruited NHs that experienced a SARS-CoV-2 outbreak in the period March to July 2020 in order to increase the likelihood of obtaining seropositive samples, hence minimizing the number of screened participants needed to obtain our sample size. The management informed the families of residents and recruited a total of 199 residents and 241 staff members interested in participating in our study.

### Sample collection

We obtained approximately 5 ml of venous blood from each participant by venipuncture in serum tubes using a 23G scalp vein set. Capillary blood was collected after briefly puncturing the top edge of the distal phalanx of the middle or ring finger using 18G safety lancets (Sarstedt, Numbrecht, Germany) onto Whatman protein saver cards (Whatman™, GE Healthcare Sciences, Cardiff, UK). During sample collections, the protein saver card of EUROIMMUN (EUROIMMUN, PerkinElmer Health Sciences Inc., Lübeck, Germany) was marketed and also evaluated on a subset of participants together with the Whatman protein saver cards. For each protein saver card, a preprinted circle was filled until saturated (i.e., blood was visible on the backside of the card). In order to avoid sampling bias, for half of the participants (i.e., those with an even participant identification number), first a circle was filled on the Whatman protein saver card, and then on the EUROIMMUN protein saver card, while for the other half of the participants (those with an odd number), the opposite was done. Typically, an average of four blood drops were needed to saturate one circle. All blood collections were done under aseptic conditions. DBS were obtained by allowing the capillary blood to air-dry on the protein saver cards for one hour at room temperature.

Serum tubes were transported to the Laboratory of Clinical Microbiology of the Ghent University Hospital (Ghent, Belgium) within six hours after sample collection. Upon arrival, serum tubes were centrifuged at 3000 g for 8 minutes, and tubes were stored at 4 °C. The following day, serum was aliquoted into new serum vials and analyzed by means of Abbott SARS-CoV-2 IgG Architect immunoassay (Abbott Laboratories). DBS selected for analysis (see further) were analyzed maximum two days after collection.

### SARS-CoV-2 IgG detection in sera by means of chemiluminescent microparticle immunoassay (CMIA)

All serum samples were analyzed for anti-nucleocapsid SARS-CoV-2 IgG serology by using the Architect i2000sr Plus system (Abbott). This system allows high-throughput screening of the sera, providing real-time information on the number of positive samples. This way, we could rapidly validate DBS for our ongoing and future studies in NHs. The Architect i2000sr Plus system uses the chemiluminescent microparticle immunoassay (CMIA)-technique to detect antibodies. After thawing the sera and vortexing briefly, the Architect system analyzes the samples automatically using a SARS-CoV-2 assay, a specific calibrator kit and a specific control kit. We used an in-house validated cut-off index of 0.9 to classify sera as positive (≥0.9) or negative (<0.9) for SARS-CoV-2 IgG antibodies.

### SARS-CoV-2 IgG detection in dried blood spots by means of ELISA

The DBS were analyzed for the presence of anti-spike (S1-antigen) SARS-CoV-2 IgG antibodies by means of ELISA (EUROIMMUN, PerkinElmer Health Sciences Inc., Lübeck, Germany), and not the Abbot CMIA, because this assay requires a volume of 150 µl, which is more than the volume of capillary blood absorbed on one circle of the DBS. From each DBS card, one circle of 6 mm diameter was cut out using a puncher and placed into the well of a sterile 96-well U-shaped microtiter plate. To avoid cross-contamination, the puncher was cleaned using a 70% alcohol solution and cotton swab in between punching. A total volume of 250 μl preheated (1 hour at 37°C) sample buffer was added to each sample well of the 96-well microtiter plate and incubated at 37 °C for 1 h. After gently mixing the eluate by means of up-and-down pipetting, a total of 100 μl of this eluate was used for ELISA, according to the manufacturer’s instructions. The ELISA was run on the automated Behring Elisa Processor III (SIEMENS AG, Munich, Germany). DBS were classified according to their antibody index optical density (OD) value as negative (<1.1) or positive (≥1.1), as recommended by the manufacturer. The borderline category was not considered.

### Sample size and analysis

The sample size was calculated using the methodology described by Buderer (1996), focusing on the sensitivity (4). Here, we hypothesized sensitivity of DBS to be lower compared to sera due to antibodies being affected by or remaining captured in the protein saver cards. Using an anticipated sensitivity of 85%, an α level of 0.05 and a precision parameter (ε) of 0.10, we needed a minimum of 49 positive sera, which were collected for both the residents and staff. The sensitivity and specificity of the DBS were calculated using the Abbott CMIA on sera from venous blood as a reference test. The 95% confidence intervals (CI) were calculated using the Wilson-Brown method (5).

To determine the optimal cut-off value for the Whatman and EUROIMMUN DBS test, we calculated Youden’s J-index statistic, and the accuracy was calculated using the area under the curve (AUC) of the receiver operating characteristic (ROC) curves.

All analyses were performed using the statistical software GraphPad Prism 6 (GraphPad Software Inc., San Diego, U.S.).

## Results

In four NHs, a total of 440 paired venous blood - Whatman DBS samples were obtained, of which 199 were from residents and 241 from staff (Figure 1). The sera from these 440 venous blood samples were screened by means of the Abbott IgG CMIA reference assay. Of these samples, 129 samples were found positive (60 residents and 69 staff). The paired Whatman DBS from these positive sera, together with 144 paired Whatman DBS samples from negative sera, were analyzed by means of ELISA. The selection of paired DBS from negative sera included 85 paired samples from residents and 59 from staff. The mean age of residents and staff of which the paired sera-Whatman DBS were analyzed was 87.8 years (range 67-100) and 42.8 years (range 19-65), respectively. A total of 80.0% and 93.8% of residents and staff, respectively, were female. Additionally, a subset of 150 EUROIMMUNE DBS samples were analyzed, of which 82 were found positive (32 residents and 50 staff) by the reference test (Figure 1).

**Figure 1.**
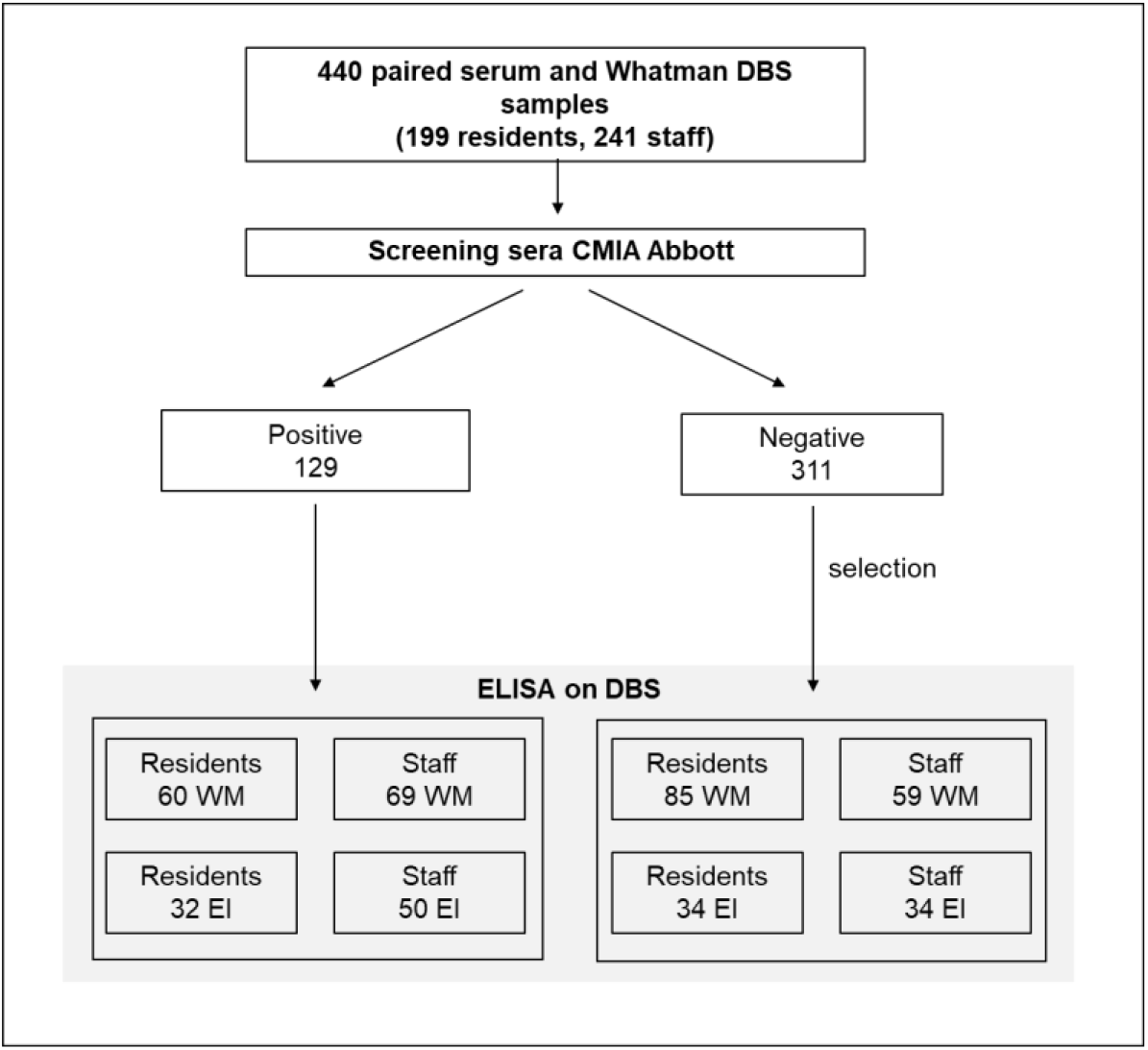
Schematic figure of the collection, screening and analysis of sera and dried blood spots. CMIA, chemiluminescent microparticle immunoassay; DBS, dried blood spot; EI, EUROIMMUN; ELISA, enzyme-linked immunosorbent assay; WM, Whatman.

The crosstabs with the number of true/false positives and negatives of the Whatman DBS and EUROIMMUN DBS in comparison with the reference test for all participants and categorized per residents and staff, is shown in Table 1. One of the two false-positive Whatman DBS test results with an optical density (OD) value of 4.39, was obtained from a participant who tested positive (RT-PCR) for SARS-CoV-2 in March 2020, making it unlikely that this DBS seropositivity was false-positive, in contrast, the reference test likely was falsely negative. The sensitivity and specificity of both the Whatman DBS and the EUROIMMUN DBS compared to the Abbott CMIA as a reference test is shown in Table 2.

**Table 1.** The number SARS-CoV-2 IgG true/false positive and true/false negative Whatman (WM) DBS and EUROIMMUN (EI) DBS in comparison to the Abbott CMIA reference test.

|  |  | Reference test (CMIA on venous blood) |  |
| --- | --- | --- | --- |
|  |  | positive | negative |
| **DBS WM total** | positive | 124 | 2 |
|  | negative | 5 | 142 |
| **DBS WM** | positive | 57 | 1 |
|  | negative | 3 | 84 |
| **DBS WM staff** | positive | 67 | 1 |
|  | negative | 2 | 58 |
| **DBS EI total** | positive | 78 | 2 |
|  | negative | 4 | 66 |
| **DBS EI** | positive | 31 | 1 |
|  | negative | 1 | 33 |
| **DBS EI staff** | positive | 47 | 1 |
|  | negative | 3 | 33 |

**Table 2.** Sensitivity and specificity of the Whatman (WM) and EUROIMMUN (EI) dried blood spot ELISA in comparison with the Abbott CMIA as the reference test^a^.

|  | **SEN (%)** | **95% CI** | **SPE (%)** | **95% CI** |
| --- | --- | --- | --- | --- |
| **DBS WM total** | 96.1 | 91.2-98.7 | 98.6 | 95.1-99.8 |
| **DBS WM** | 95.0 | 86.3-98.6 | 98.8 | 93.6-99.9 |
| **DBS WM staff** | 97.1 | 90.0-99.5 | 98.3 | 91.0-99.1 |
| **DBS EI total** | 95.1 | 88.1-98.1 | 97.1 | 89.9-99.5 |
| **DBS EI residents** | 96.7 | 84.3-99.8 | 97.1 | 85.1-99.9 |
| **DBS EI staff** | 94.0 | 83.8-98.4 | 97.1 | 85.1-99.9 |

<sup>a</sup>CMIA, chemiluminescent microparticle immunoassay; CI, confidence interval; DBS, dried blood spot; EI, EUROIMMUN; SEN, sensitivity; SPE, specificity; WM, Whatman.

The scatter plot of the index values of the reference test (Abbot CMIA) and the OD values of the ELISA Whatman DBS is shown in Figure 2. The Pearson correlation coefficient for the reference test and the Whatman DBS was 0.80 (95% CI, 0.75 to 0.84) and significant (p<0.0001). The Pearson correlation coefficient for the reference test and the EUROIMMUN DBS was 0.78 (95% CI, 0.73 to 0.82) and significant (p<0.0001) (scatterplot similar to reference test versus Whatman DBS, data not shown). A scatter plot showing the correlation between the optical density (OD) values from the 150 paired Whatman and EUROIMMUN DBS is depicted in Figure 2. The Pearson correlation coefficient was found to be 0.970 (95% CI, 0.959 - 0.978) and significant (p<0.0001). Of one paired sample, the Whatman DBS showed a positive result (OD 1.41), but not the EUROIMMUN DBS (OD 1.02). However, the latter is classified as borderline when following the manufacturer’s guidelines. This category was not considered in the current analysis and classified as negative.

**Figure 2.**
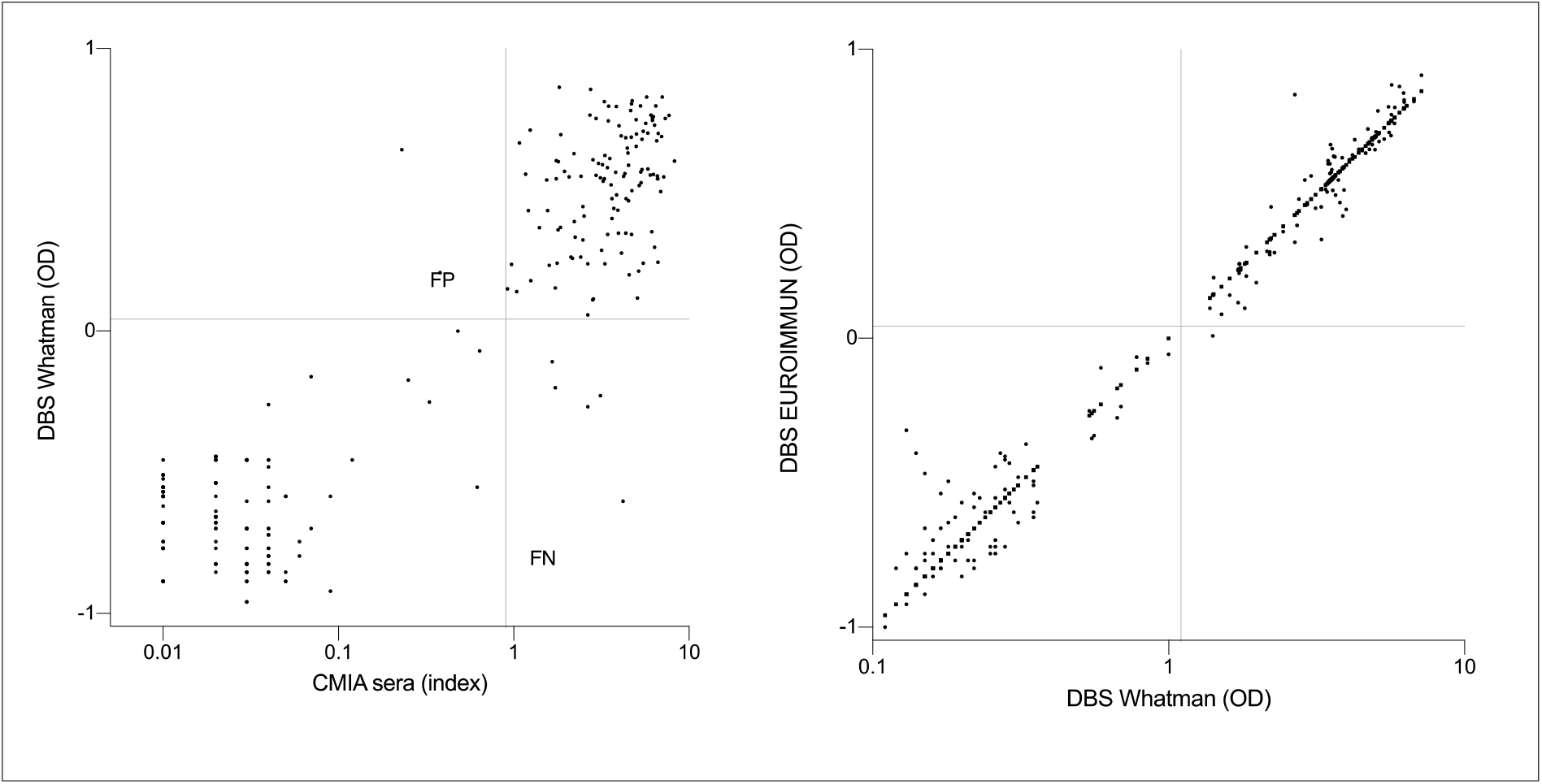
Scatter plot of paired sera and Whatman DBS (left) and Whatman DBS and EUROIMMUN DBS (right) optical density/index values. Axes represent the log10 of the optical density and index values. DBS, dried blood spot; FN, false negatives; FP, false positives; OD, optical density. The grey horizontal and vertical lines represent cutoff lines defining positive samples.

To verify if the ELISA Whatman and EUROIMMUN DBS cutoff could be optimized, we performed a ROC analysis. The ROC curves are shown in Figure 3. The area under the curve was 0.999 for both Whatman and EUROIMMUN DBS (95% CI, 0.999 to 1.000) (p<0.0001). The optimal cut-off point for Whatman DBS was found to be 1.14, which is almost exactly the value of 1.1 proposed by the manufacturer of the ELISA assay, and did not improve sensitivity or specificity. The optimal cut-off point for EUROIMMUN DBS was found to be 1.02 and applying this cutoff improved both sensitivity (from 95.1% to 96.4%) and specificity (from 97.1% to 98.5%).

**Figure 3.**
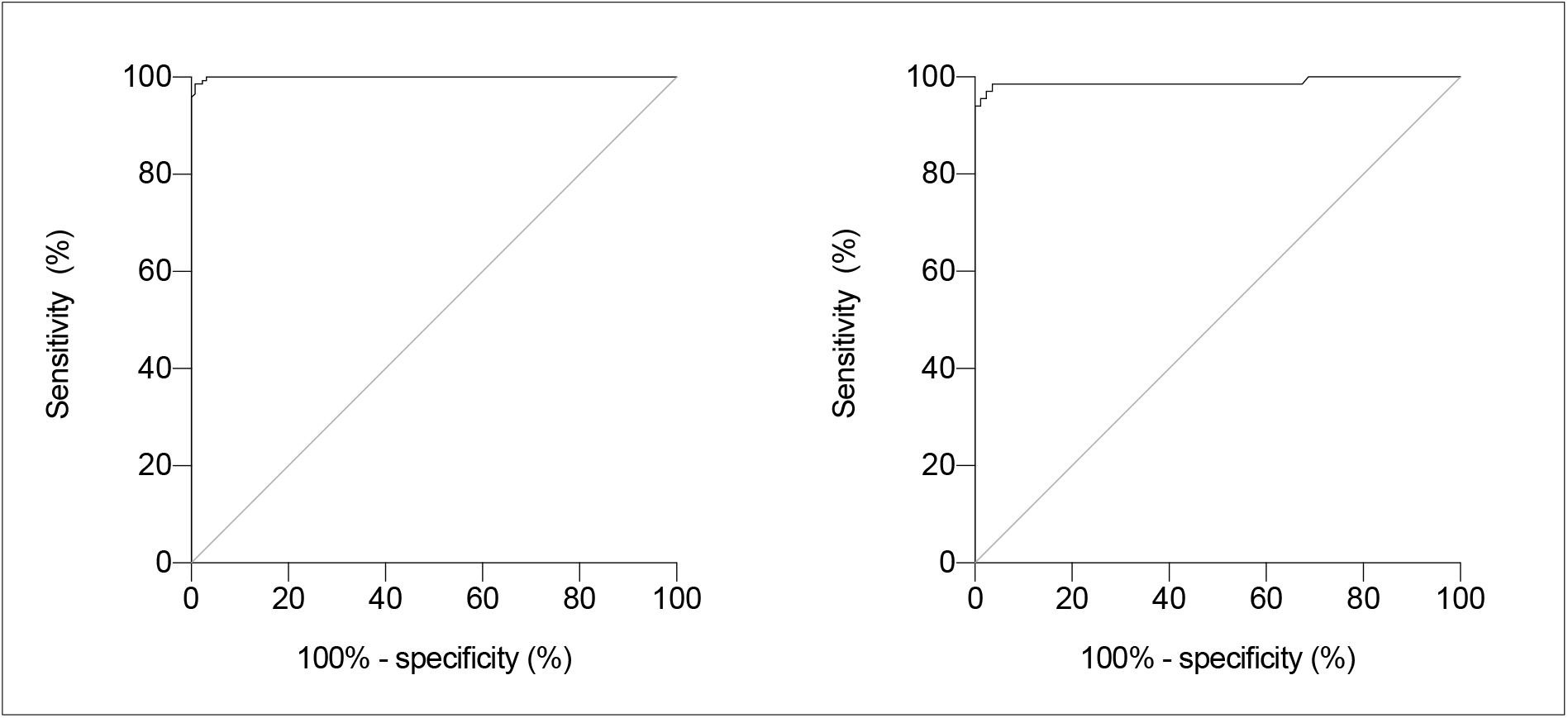
Receiver operating characteristics (ROC) curve for the evaluation of the Whatman DBS ELISA (left) and EUROIMMUN (right) compared to the reference assay. The area under the curve is 0.999 for both Whatman and EUROIMMUN DBS (95% CI, 0.999 to 1.000) (p<0.0001).

## Discussion

In the present study, we evaluated the use of DBS for detection of SARS-CoV-2 antibodies in residents and staff from nursing homes. To the best of our knowledge, we report the largest clinical DBS validation study, comparing two different DBS samples (Whatman and EUROIMMUN) in a key target population in the current SARS-CoV-2 pandemic, the elderly persons residing in NH.

Overall, we demonstrated a high sensitivity (range 95.0-97.1%) and specificity (range 97.1-98.8%) for DBS compared to sera. Adjustment of the manufacturer recommended cut-off from 1.1 to 1.02 resulted in a slight improvement of sensitivity and specificity for EUROIMMUN DBS only. We found no significant differences in sensitivity or specificity of the DBS between residents and staff, for both Whatman and EUROIMMUN DBS, as demonstrated by the largely overlapping confidence intervals. Furthermore, protein saver cards of Whatman and EUROIMMUN yielded results that were nearly in perfect agreement.

Several other studies in different populations, such as health care workers, key workers, athletes and children have evaluated DBS sampling for SAR-CoV-2 antibody testing (6–14). In these studies, similar test characteristics were reported with a sensitivity ranging from 89 up to 100% and a specificity of 100% (7, 10, 11), or, reported a nearly perfect agreement in SARS-CoV-2 antibody detection between DBS samples and paired venous blood samples (6, 9, 12–14). However, some studies were limited by the lack of serum serology as a reference test (7, 8) or the rather limited sample size (i.e., <50 positives as assessed by the reference test) (6, 7, 11, 12).

Our findings show an excellent diagnostic performance of DBS, both in the elderly and staff from nursing homes. This supports the use of DBS in large-scale SARS-CoV-2 serosurveillance studies as a valuable alternative to venipuncture, especially in the elderly, where venous blood can be challenging to obtain. In comparison to serological rapid tests, which can also be implemented in large-scale SARS-CoV-2 sero-epidemiological studies, DBS offer higher sensitivity, specificity and the possibility to (semi-)quantitatively asses the antibody response (15). Moreover, DBS have the advantage to collect and store the sample for multiple analyses. In this way, different assays can be used on the same sample, such as antibody assay directed against different antigens. Yet, DBS have similar advantages as these rapid tests, as they alleviate the need for health care professionals during sampling and complex shipment and storage.

Especially since the implementation of newly developed SARS-CoV-2 vaccines, serology studies are of increasing importance to fill existing research gaps. Until now, the stability and duration of the antibody response upon vaccination is unknown, however, is important to determine if annual boosting is needed. Secondly, a correct antibody response cut-off value that refers to clinical immunity against SARS-CoV-2 is lacking. Additionally, it is of crucial importance to assess the effectiveness of the vaccines among the general population, which can differentiate from what is measured in standardized clinical trials. In this context, the non-invasiveness of the DBS, minimal logistic constraints, excellent test characteristics and the possibility for quantitative assessment bring added value in the conduct of serosurveillance studies.

Our study was limited by the use of CMIA on venous blood samples that assessed nucleocapsid antibodies, and not spike antibodies, which were detected by means of ELISA in DBS. Conflicting results are available concerning the degree of persistence between antibodies directed against the nucleocapsid and spike antigen, however, it is suggested that IgG antibody levels directed against the nucleocapsid antigen wane more rapidly post-infection than those against the spike antigen (16). Nevertheless, this should not impact the sensitivity analysis in the current results, as nucleocapsid antibodies were detected as the reference.

## Data Availability

Not applicable

## Acknowledgements

The authors wish to thank all residents and their families, staff and management from the nursing homes that participated in the study.

